# Antimicrobial Susceptibility Pattern and Detection of Extended-Spectrum Beta-Lactamase (blaCTX-M) Gene in *Escherichia coli* from Urinary Tract Infections at the University Teaching Hospital in Lusaka, Zambia

**DOI:** 10.1101/2020.05.16.20103705

**Authors:** Emmanuel Chirwa, Georgina Mulundu, Kunda Ndashe, Kalo Kanongesha, Wezi Kachinda, Kaziwe Simpokolwe, Bernard Mudenda Hang’ombe

## Abstract

**Background:** Urinary tract infections caused by *Extended Spectrum Beta-Lactamase* producing *Escherichia coli* are increasing globally and yet treatment still remains a challenge due to antibiotic resistance of the causative agent.

**Objectives:** The aim of the study was to determine the antimicrobial susceptibility pattern and detect the presence of blaCTX-M gene in *Escherichia coli* isolated from urinary tract infection patients at the University Teaching Hospital, Lusaka, Zambia.

**Methodology:** This was a cross sectional study that involved collection of urine samples from patients who were diagnosed with urinary tract infections. The samples were cultured on MacConkey agar complemented with cefotaxime and Polymerase Chain Reaction was performed to confirm the *Extended Spectrum Beta-Lactamase* producers by detecting the CTX-M gene. Antimicrobial susceptibility tests were conducted using standard methods.

**Results:** A total of 327 urine samples were cultured and 15 (4.6%) of these samples were positive ESBL producers. The isolates showed complete resistance to ampicillin and cotrimoxazole.

**Conclusion:** Multi drug resistant *Extended Spectrum Beta-Lactamase* producing *Escherichia coli* was detected in 4.6 % of UTI patients at the University Teaching Hospital.

## INTRODUCTION

Urinary tract infections (UTIs) are some of the most common bacterial infections affecting 150 million people each year globally (Stamm and Norrby, 2001). In Africa, two studies reported that the prevalence of UTIs was 0.7°C and 4.5°C in Algeria and Senegal, respectively, (Atif et al., 2006; Dia et al., 2008), while a retrospective study from Nigeria reported a frequency of 12.3°C (Jombo et al., 2006). A study conducted in Zambia reported a prevalence of 33°C of UTIs among paediatrics at the University Teaching Hospital in Lusaka district [5].

The spectrum of bacteria causing UTIs include *Escherichia coli* (*E. Coli*), *Citrobacter* spp, *Enterobacter aerogenes, Pseudomonas aeruginosa, Proteus vulgaris*, and *Klebsiella* spp, whereas *Staphylococcus* spp, and *Salmonella* spp. are found rarely (Abujnah et al., 2015; Davies and Davies, 2010). Chisanga and colleagues reported that *E. coli, Klebsiella* spp, *Proteus* spp and *Staphylococcus saprophyticus* were isolated from patients with UTIs in Zambia (Chisanga et al., 2017). Therefore *E. coli* and *Klebsiella* spp remain the commonest uropathogens that cause severe infections. These bacteria have the ability to produce *Extended Spectrum Beta-Lactamase* (ESBL), which are plasmid-borne enzymes that confer multiple drug resistance hence making UTIs difficult to treat (Lautenbach et al., 2001; Rupp and Fey, 2003). The ESBL producing bacteria have shown resistance to fluoroquinolones, aminoglycosides, and fourth-generation cephalosporins, (Kariuki et al., 2007; Lautenbach et al., 2001).

*Extended Spectrum Beta-Lactamases* mainly include TEM, SHV, CTX-M, VEB, and GES enzymes (Bonnet, 2004) and the highest number of variants corresponds to the CTX-M family (Cantón and Coque, 2006). CTX-M *beta lactamases* produced by *Escherichia coli* have emerged worldwide as an important cause of infections and this has been described as the “the CTX-M pandemic” (Cantón and Coque, 2006). During the past two decades these enzymes have been reported as the commonest type of ESBL found in most areas of the world (Bonnet, 2004). In Africa, *E.coli* producing CTX-M *beta lactamase* have been identified in several counties which include Algeria, Tunisia, Cameroon, Tanzania and the Central African Republic(Bercion et al., 2009; Blomberg et al., 2005; Gangouó-Pióboji et al., 2005; Ramdani-Bouguessa et al., 2006; Sallem et al., 2012). The proportion of ESBL-producing bacteria causing common infections in all regions of the world is high, making antibiotic resistance due to ESBL being a major global public health problem.

This study was undertaken in view of “the CTX-M pandemic” globally as reported by other researchers. The aim of the study was to determine the antimicrobial susceptibility pattern and detect the presence of blaCTX-M gene in *E coli* isolated from UTI patients at the University Teaching Hospital, Lusaka, Zambia.

## MATERIALS AND METHODS

### Study Area

The study was conducted at the University Teaching Hospital (UTH), Lusaka, Zambia. The urine samples were collected from patients that reported to Urology, outpatient and Maternity departments and were diagnosed with UTI.

### Study population

The urine samples of 327 patients, both in patients and out patients who attended the urology department of UTH and had evidence of UTI determined by treating physicians were included in the study. Patients on antibiotic therapy were excluded from the study. Ethical clearance was obtained from the University of Zambia Biomedical Research Ethics Committee (Ref. 007–01–17) and a written consent and assent form was sought from each patient or bed-sider.

### Sample Collection

Clean catch midstream urine was collected from each patient into 20mL sterile universal container. The specimen were labelled and transported to the bacteriology laboratory at the University Teaching Hospital, with 6 hours of collection. In each container boric acid (0.1g/10ml of urine) was added to prevent the growth of bacteria in the urine sample.

### Isolation of ESBL Escherichia coli

Initial screening was conducted by inoculating each sample on MacConkey agar (Oxoid, UK) supplemented with cefotaxime at a screening concentration of 2μg/ml and incubated at 37°C for 24 hours. The positive cultures were then sub cultured on MacConkey agar (Oxoid, UK) and pure colonies were then obtained. Identification of bacterial isolates was done on the basis of their cultural and biochemical characteristics.

### Molecular detection of blaCTX-M gene

For detection of blaCTX-M gene, the isolates were cultured in Brain Heart Infusion, (Himedia, India) at 37°C for 18 hours. After incubation, 1ml of bacterial suspension was centrifuged at 5800 xg for 5 minutes, thereafter, the supernatant was discarded. The remaining cell pellet was washed with 500μl of normal saline, centrifuged at 13000 xg for 5 minutes and the supernatant discarded. After washing, 500μl of TE buffer (pH 8.0) was added to the cell pellet and then heat treated until boiling, then immediately transferred to ice for 10 minutes. The cell debris was removed after centrifuging at 13000 xg for 5 minutes, while the supernatant was transferred into a new microfuge tube and stored at –20°C until use (Paterson and Bonomo, 2005).

The master mix per reaction tube was made with 5μl of Phusion Flash, 2μl sterile water, 1μl of reverse primer, 1μl of forward primer and 1μl of DNA template, giving total volume of 10μl per reaction tube, which was mixed using a vortex mixer. The PCR primers used for the detection of blaCTX-M are indicated in Table 1. The PCR conditions were set as 98% for 30 seconds, 98% for 0 seconds, 60% for 5 seconds(35 cycles), 72% for 15 seconds, 72% for 2 minutes, and holding at 4% infinitely. After PCR, the products were visualized on a trans-illuminator machine following staining with ethidium bromide [20].

**Table 1:**
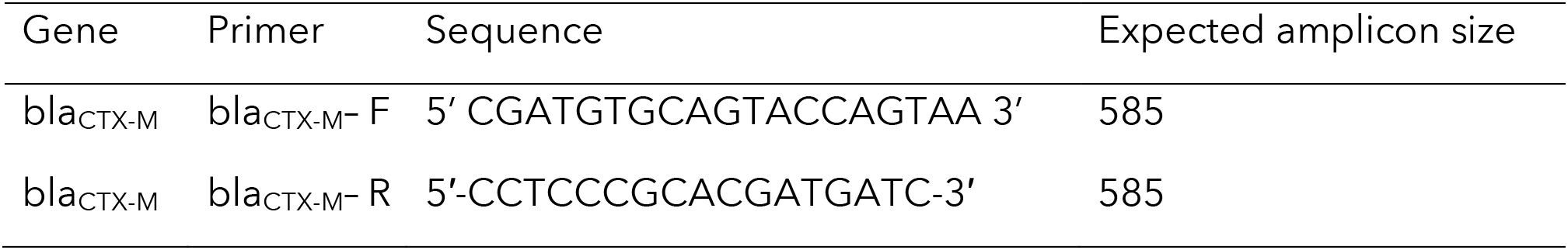
blaCTX-M Primers sequences

#### Antibiotic Susceptibility Testing

The blaCTX-M positive isolates were tested for antimicrobial susceptibility testing by the standard Kirby Bauer’s disc diffusion method. Standard inoculums adjusted to 0.5 McFarland was swabbed on Mueller Hinton agar and was allowed to soak for 2 to 5 minutes. After that antibiotic disks were placed on the surface of media and pressed gently. Mueller Hinton agar plates were then incubated at 37% for 24th. After 24h the inhibition zones were measured and interpreted by the recommendations of clinical and laboratory standards [21]. The following standard antibiotic discs were used for the isolates, Ampicillin (A), Cotrimoxazole (Co), Tetracycline (TE), Gentamycin (G), Nitrofurantoin (NIT), Chloramphenicol (C), Nalidixic acid (NA), Cephotaxin (CTX), Norfloxacin (NX) and Ciprofloxacin (CIP).

## RESULTS

The overall prevalence of ESBL E. coli in UTI patients was found to be 4.6% (15/327), this was confirmed by culturing urine samples on MacConkey agar supplemented with cefotaxime. Of the 15 ESBL *E. coli* positive samples, blaCTX-M gene was detected in 14 samples [figure 1].

**Figure 1:**
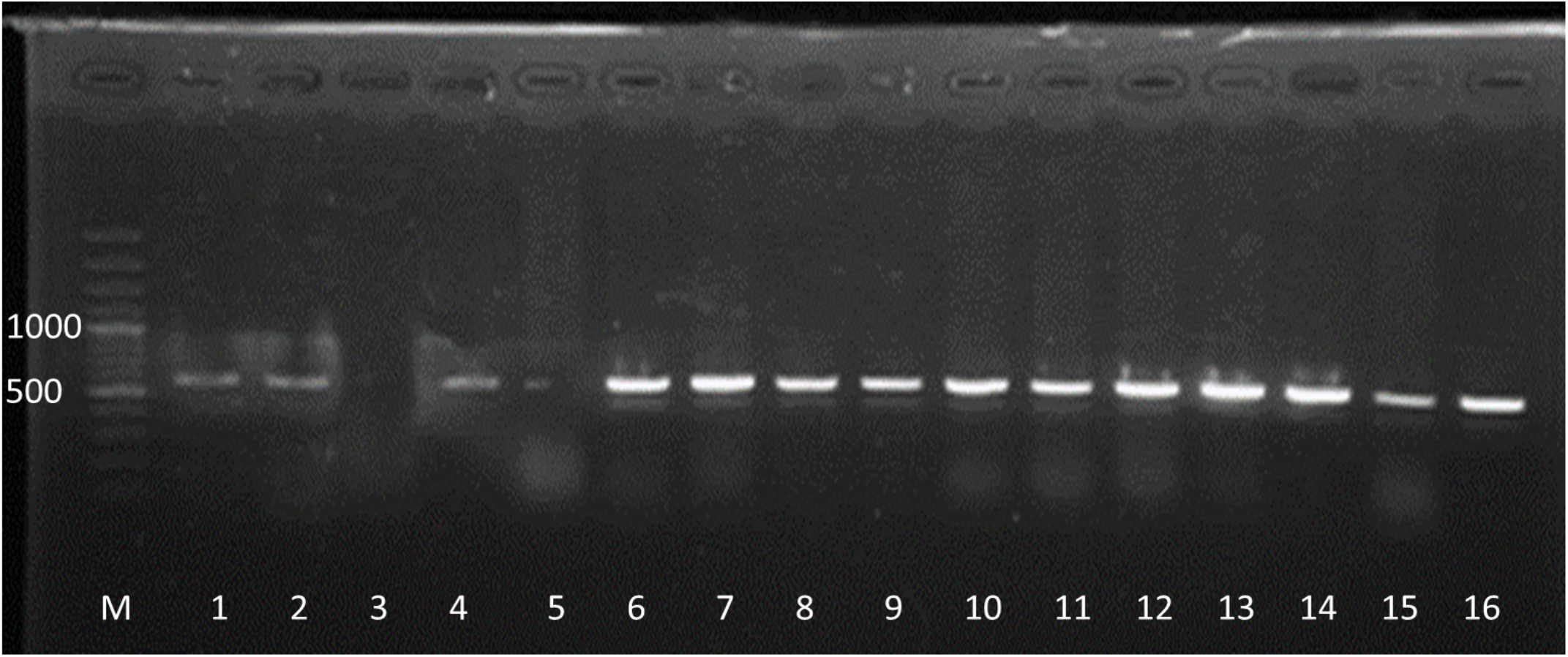
Gel electrophoresis picture for blaCTX-M gene in E coli from UTI. Lane M; DNA ladder (100bp), Lane 1 to 15 ESBL E coli positive samples, and Lane 16 the positive control.

Antibiogram by Kirby and Bauer’s method was carried out for all the 15 identified isolates of *E. coli* and a total of 10 antibiotics were used. The percentage of resistance clearly indicates that ESBL producers are totally not responding to Ampicillin (100%), Cotrimoxazole (100%), Cephotaxin (87%), Tetracycline (53%), Nalidixic acid (47%), Norfloxacin (47%), Ciprofloxacin (47%), Chloramphenicol (40%), Gentamycin (13%) and Nitrofurantoin (0%) [Figure 2].

**Figure 2:**
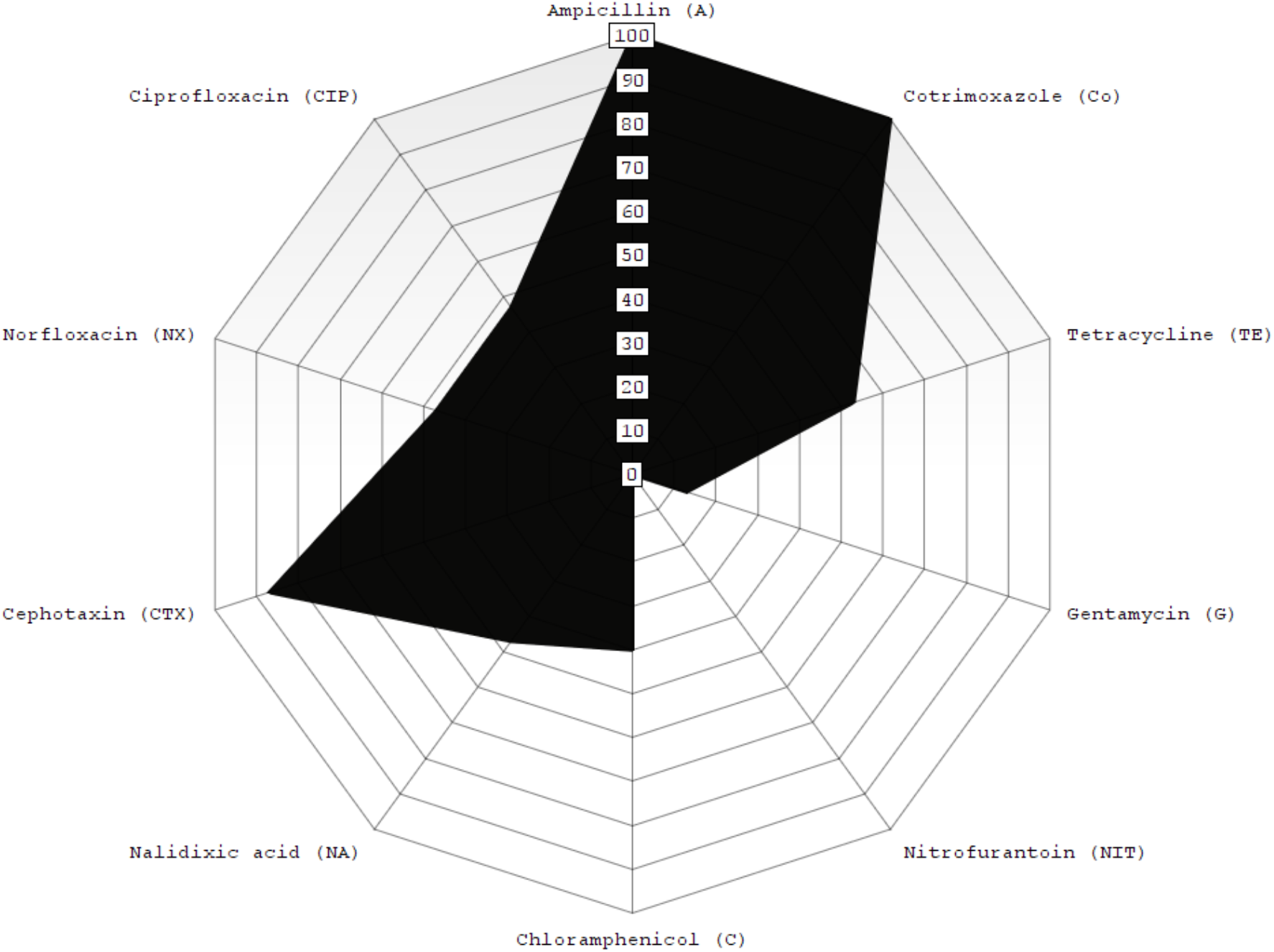
Antimicrobial resistance pattern of ESBL E. coli isolates detected with the CTXM gene. The radar chart shows percentage resistance of the antibiotics used in the study, Ampicillin (100%), Cotrimoxazole (100%), Cephotaxin (87%), Tetracycline (53%), Nalidixic acid (47%), Norfloxacin (47%), Ciprofloxacin (47%), Chloramphenicol (40%), Gentamycin (13%) and Nitrofurantoin (0%).

## DISCUSSION

The ESBL *E. coli* proportion estimate of patients clinically diagnosed with UTI in this study was 4.6 %. The results of this study indicate lower estimates than those reported in other African countries such as Tunisia (7.3%), Cameroon (12%), Algeria (20%) and Tanzania (25%) (Blomberg et al., 2005; Gangouó-Pióboji et al., 2005; Ramdani-Bouguessa et al., 2006; Sallem et al., 2012). Furthermore, in resource-rich countries such as Germany the average ESBL proportion estimates reported in a nationwide hospital survey for 2012 were between 10 to 15 % (Leistner et al., 2015). In the same year 2012, in United States of America the ESBL proportion estimates ranged between 4 to 12 % (Castanheira et al., 2014). It is important to note that the present study focused on both nosocomial and community acquired infections while the other studies were concerned with nosocomial infections only. The differences in ESBL proportion estimates maybe attributed to variations in populations surveyed, difference in clinical diagnostic methods for UTIs, or by the levels of antibiotic use in the community.

In the present study, genes coding for CTX-M were detected in 93% of the samples that were positive for ESBL *E. coli* on culture. Currently, CTX-M enzymes include more than 80 different enzymes that are clustered into 5 groups based on their amino acid sequences and the include CTX-M-1, –2, –8, –9 and –25 (Paterson and Bonomo, 2005). However, sequencing was not done to discriminate between the 5 CTX-M groups. The detection of CTX-M gene in 4.3% and 93% of clinically diagnosed UTI cases and ESBL *E. coli* isolates, respectively, is indication of growing problem in UTIs being caused by drug resistant bacteria. The study collected urine samples from both in-patients and out-patients at the University Teaching Hospital, therefore indicating the presence of CTX-M gene in both nosocomial and community UTI infections. The spread of CTX-M gene in the community has already been described through studies conducted in industrialized countries such as Canada (Pitout et al., 2004), France (Arpin et al., 2003), and the United Kingdom (Woodford et al., 2004). Although not determined in this study, risk factors for acquiring community-onset infection due of CTX-M producing *E. coli* include repeat UTIs, underlying renal pathologies, previous hospitalisation, nursing home residents, diabetes mellitus, underlying liver pathology and international travel (Laupland et al., 2008; Paterson and Bonomo, 2005; Rodríguez-Baño et al., 2008).

Multidrug resistance was recorded in all the *ESBL E. coli* isolates in this study. The resistance was very high to Ampicillin (100%), Cotrimoxazole (100%) and Cephotaxin (87%). The resistance patterns were extended even to non-beta-lactam antibiotics such as nalidixic acid (47%), tetracycline (53%), norfloxacin (47%), ciprofloxacin (47%) and chloramphenicol (40%). Quinolones have been used to treat complicated UTIs associated with ESBL-producing organisms without in vitro resistance to the drug (Paterson and Bonomo, 2005). However, recent studies associate ESBL production with fluoroquinolone resistance (Rupp and Fey, 2003), and in this study, resistance was high for ciprofloxacin (47%), norfloxacin (47%) and nalidixic acid (47%). The resistance to both beta-lactam antibiotics and other classes of antibiotics in this study suggests the presence of other antimicrobial resistance genes in *E. coli* isolates in addition to ESBL genes, leading to multidrug resistance (Paterson, 2006; Rao et al., 2014). The observed multidrug resistance in ESBL producing isolates has an implication on treatment, limiting the available options for curative therapy for UTIs. Globally, there has been increased resistance to commonly used antibiotics. Self-medication and lack of completion of antibiotic treatment has been recognised as some of the contributing factors to antibiotic resistance (Mshana et al., 2013).

## CONCLUSION

The prevalence of extended spectrum beta lactamase –producing Escherichia coli among urinary tract infections at University Teaching Hospital, Zambia are 4.5 %. The CTX-M gene was detected in 93% of ESBL E. coli isolates. Furthermore, ESBL *E. coli* isolates showed multi drug resistance

With complete resistance to ampicillin and Cotrimoxazole but was susceptible to nitrofurantoin.

## Data Availability

All data obtained in the study is available to readers and reviewer is available within the manuscript

## ACKNOWLEDGEMENT

We would like to recognize that this was a study undertaken by Emmanuel Chirwa as partial fulfilment for Master of Science Medical Microbiology of the University of Zambia. We wish to express our gratitude to the staff at the University Teaching Hospital, Bacteriology Laboratory and School of Veterinary Medicine, Bacteriology and Molecular Diagnostics Laboratories for technical help rendered during bench work sessions of the research. Finally, we are indebted to the patients of University Teaching Hospital that allowed us to collect samples to conduct the research, to them we say thank you.

